# Cognitive impairment is a common comorbidity in COVID-19 deceased patients. A hospital-based retrospective cohort study

**DOI:** 10.1101/2020.06.08.20125872

**Authors:** Paloma Martín-Jiménez, Mariana I. Muñoz-García, David Seoane, Lucas Roca-Rodríguez, Ana García-Reyne, Antonio Lalueza, Guillermo Maestro, Dolores Folgueira, Víctor A. Blanco-Palmero, Alejandro Herrero-San Martín, Sara Llamas-Velasco, David A. Pérez-Martínez, Marta González-Sánchez, Alberto Villarejo-Galende

## Abstract

**Introduction:** Little is known about the relation of cognitive impairment (CI) to COVID-19 mortality. Here, we analyse the frequency of CI in deceased COVID-19 patients.

**Methods:** We included 477 adult cases that died after admission from March 1 to March 31, 2020: 281 with confirmed COVID-19, 58 probable COVID-19, and 138 who died of other causes.

**Results:** The number of comorbidities was high in the confirmed COVID-19, and CI was common (30%: 21.1% dementia; 8.9% mild cognitive impairment). Subjects with CI were older, more lived in nursing homes and had shorter times from symptom onset to death than those without CI. COVID-19 patients with CI were rarely admitted to the ICU and fewer received non-invasive mechanical ventilation, but palliative care was provided more often.

**Conclusions:** Dementia is a frequent comorbidity in COVID-19 deceased patients. The burden of COVID-19 in the dementia community will be high.

## INTRODUCTION

On January 7, 2020, Chinese investigators described a novel coronavirus (SARS-CoV-2) as the cause of an outbreak of acute respiratory syndrome in Wuhan, China (1). Since then, the situation escalated rapidly, with an increasing number of cases in many countries. COVID-19 has had a substantial case fatality rate and a huge impact on society and healthcare. Previous reports have described some vulnerable demographic groups associated with death from COVID-19, like male sex, older age, or patients with comorbidities like hypertension, diabetes, cardiovascular disease or cancer (2). Information about neurological comorbidities in COVID-19 is scarce until now. Recently, it has been pointed out that patients with dementia may have an increased risk of COVID-19, because they can have difficulties to remember safeguard measures, and many of them live in nursing homes where the disease can spread rapidly (3). Nevertheless, the first clinical studies that analysed comorbidities and death in COVID-19 did not include dementia or cognitive impairment (CI) (4–9). In the present study, we aim to analyse the frequency of CI and other neurological comorbidities in deceased patients with COVID-19 in our hospital.

## METHODS

In this retrospective study, we reviewed all patients older than 16 years who died after admission, from March 1, 2020, to March 31, 2020, at Hospital Universitario 12 de Octubre, a tertiary hospital situated in Madrid, Spain. Since 2011, the hospital uses integrated electronic medical records that include all medical encounters and diagnoses. All data from deceased patients were manually extracted from these records using a standardized data collection form. Diagnoses of CI had been made by neurologists, psychiatrists, geriatricians, internal medicine doctors or general practitioners.

Patients were included in three groups according to COVID-19 status, following the case definition of the European Centre for Diseases Prevention and control (10):

- Confirmed COVID-19 related death. These cases were confirmed through reverse transcriptase–polymerase chain reaction (RT-PCR) assays performed on nasopharyngeal swabs.
- Probable COVID-19 related death. These cases had clinical, laboratory and radiological features consistent with COVID-19, but the RT-PCR was negative or could not be performed.
- Death not related to COVID-19: patients who died from other causes.

The study was reviewed and approved by the local ethics committee / institutional review board (EC/IRB).

Statistical analyses were undertaken using STATA software 14.0 (StataCorp, College Station, TX). Continuous variables were presented as median and interquartile range (IQR), and categorical variables as total number (n) and percentage (%). First, we compared two groups, those with confirmed COVID-19 and those with other causes of death. For other analyses we used only COVID-19 confirmed cases, comparing those with and without CI. Categorical variables were compared using the χ2 test. Means for continuous variables were compared using independent group *t* tests. All tests were two-sided, and the significance level was set at p<0.05.

## RESULTS

The number of admissions during the study period was 4,156 patients, and 1,970 were COVID-19 cases. The total number of deaths was 477. Of them, 281 died from confirmed COVID-19, 58 probable COVID-19, and 138 from other causes.

Demographic characteristics and comorbidities of the three groups are shown in **Table 1**. Median age was higher in both COVID-19 groups, and the percentage of males was significantly higher in the COVID-19 confirmed group (62.3% *vs*. 49.3%, *p*<0.01). Comorbidities were common in the three groups. The proportion of subjects with CI was similar, around 30% in each of them.

**Table 1.**
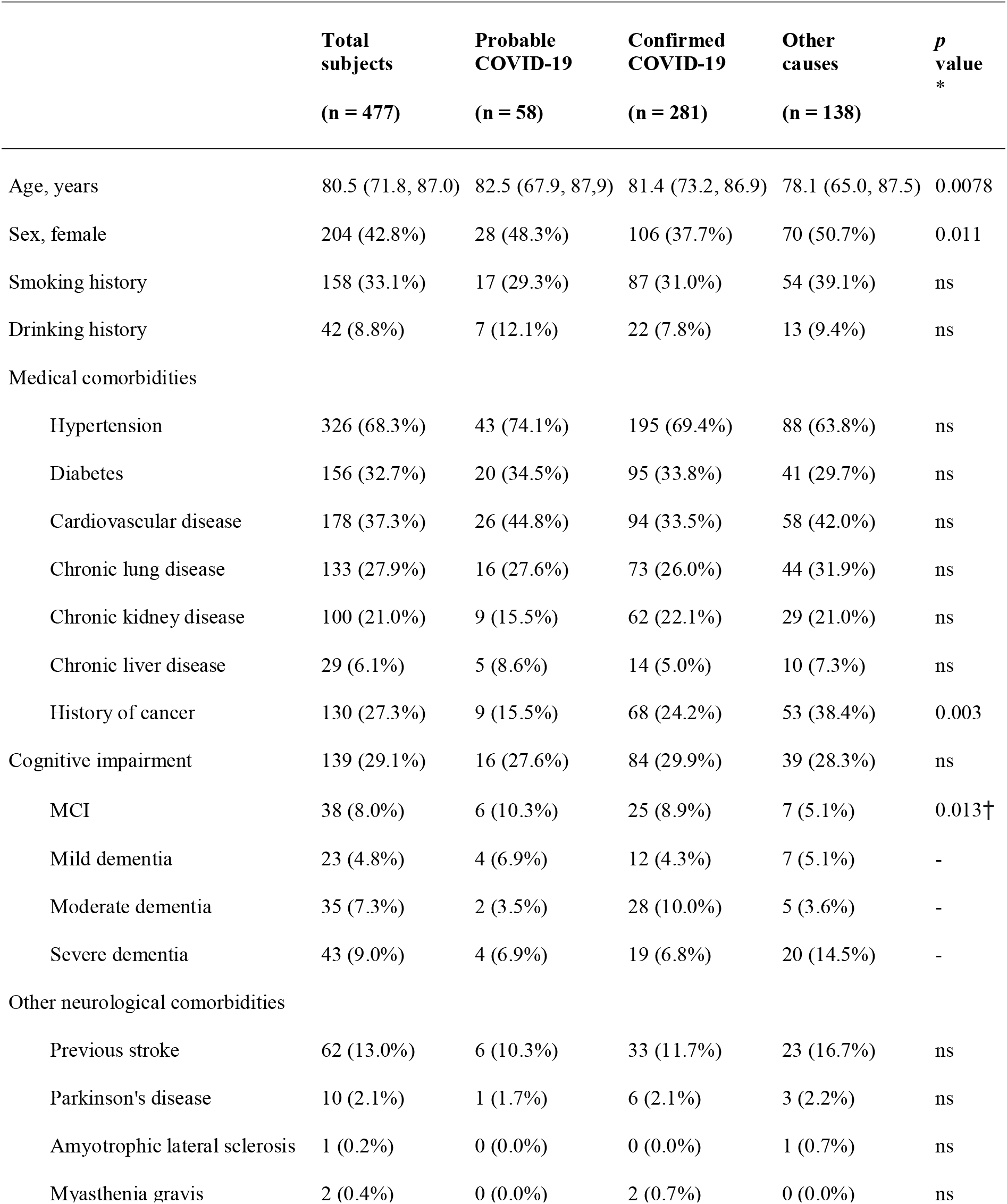
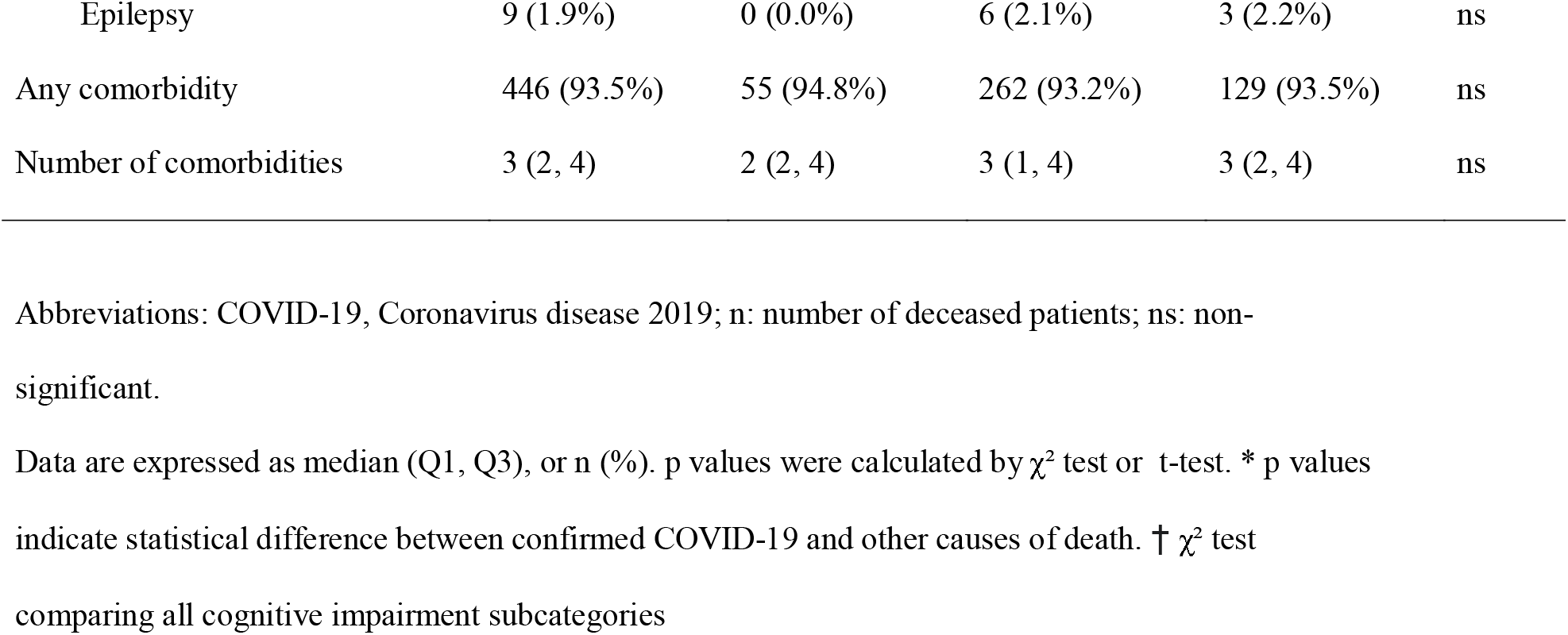
Clinical and demographic characteristics of deceased patients with COVID-19 or other causes.

|  | **Total**<br>**subjects**<br>**(n = 477)** | **Probable**<br>**COVID-19**<br>**(n = 58)** | **Confirmed**<br>**COVID-19**<br>**(n = 281)** | **Other**<br>**causes**<br>**(n = 138)** | **p**<br>**value**<br>**\*** |
| --- | --- | --- | --- | --- | --- |
| Age, years | 80.5 (71.8, 87.0) | 82.5 (67.9, 87.9) | 81.4 (73.2, 86.9) | 78.1 (65.0, 87.5) | 0.0078 |
| Sex, female | 204 (42.8%) | 28 (48.3%) | 106 (37.7%) | 70 (50.7%) | 0.011 |
| Smoking history | 158 (33.1%) | 17 (29.3%) | 87 (31.0%) | 54 (39.1%) | ns |
| Drinking history | 42 (8.8%) | 7 (12.1%) | 22 (7.8%) | 13 (9.4%) | ns |
| **Medical comorbidities** |  |  |  |  |  |
| Hypertension | 326 (68.3%) | 43 (74.1%) | 195 (69.4%) | 88 (63.8%) | ns |
| Diabetes | 156 (32.7%) | 20 (34.5%) | 95 (33.8%) | 41 (29.7%) | ns |
| Cardiovascular disease | 178 (37.3%) | 26 (44.8%) | 94 (33.5%) | 58 (42.0%) | ns |
| Chronic lung disease | 133 (27.9%) | 16 (27.6%) | 73 (26.0%) | 44 (31.9%) | ns |
| Chronic kidney disease | 100 (21.0%) | 9 (15.5%) | 62 (22.1%) | 29 (21.0%) | ns |
| Chronic liver disease | 29 (6.1%) | 5 (8.6%) | 14 (5.0%) | 10 (7.3%) | ns |
| History of cancer | 130 (27.3%) | 9 (15.5%) | 68 (24.2%) | 53 (38.4%) | 0.003 |
| Cognitive impairment | 139 (29.1%) | 16 (27.6%) | 84 (29.9%) | 39 (28.3%) | ns |
| MCI | 38 (8.0%) | 6 (10.3%) | 25 (8.9%) | 7 (5.1%) | 0.013† |
| Mild dementia | 23 (4.8%) | 4 (6.9%) | 12 (4.3%) | 7 (5.1%) | - |
| Moderate dementia | 35 (7.3%) | 2 (3.5%) | 28 (10.0%) | 5 (3.6%) | - |
| Severe dementia | 43 (9.0%) | 4 (6.9%) | 19 (6.8%) | 20 (14.5%) | - |
| **Other neurological comorbidities** |  |  |  |  |  |
| Previous stroke | 62 (13.0%) | 6 (10.3%) | 33 (11.7%) | 23 (16.7%) | ns |
| Parkinson's disease | 10 (2.1%) | 1 (1.7%) | 6 (2.1%) | 3 (2.2%) | ns |
| Amyotrophic lateral sclerosis | 1 (0.2%) | 0 (0.0%) | 0 (0.0%) | 1 (0.7%) | ns |
| Myasthenia gravis | 2 (0.4%) | 0 (0.0%) | 2 (0.7%) | 0 (0.0%) | ns |
| Epilepsy | 9 (1.9%) | 0 (0.0%) | 6 (2.1%) | 3 (2.2%) | ns |
| Any comorbidity | 446 (93.5%) | 55 (94.8%) | 262 (93.2%) | 129 (93.5%) | ns |
| Number of comorbidities | 3 (2, 4) | 2 (2, 4) | 3 (1, 4) | 3 (2, 4) | ns |

Abbreviations: COVID-19, Coronavirus disease 2019; n: number of deceased patients; ns: non-significant.
Data are expressed as median (Q1, Q3), or n (%). p values were calculated by $\chi^2$ test or t-test. \* p values indicate statistical difference between confirmed COVID-19 and other causes of death. † $\chi^2$ test comparing all cognitive impairment subcategories

Focusing on the confirmed COVID-19 group, CI was the fourth most frequent comorbidity, after hypertension (69.4%), diabetes (33.8%) and cardiovascular disease (33.5%). **Table 2** analyses separately those with and without CI. Subjects with CI were older, more of them lived in nursing homes and had with less frequency chronic lung disease. In this group, 21.1% had dementia (mild: 4.3%; moderate: 10.0%; severe: 6.8%), and 8.9% mild cognitive impairment. The most common diagnoses of CI in the COVID-19 confirmed group were Alzheimer’s disease (9.3% of all patients), mixed (7.2%) or vascular CI (4.8%). History of previous stroke or Parkinson’s disease was more frequent in those with CI. Times from symptom onset to ED and from symptom onset to death were lower in subjects with CI, and they had fever upon arrival at the ED with less frequency (32.5% *vs*. 52.2%, *p*=0.004). Encephalopathy was the most common neurological complication in both groups, and it was more common in patients with CI (32.1% *vs*. 14.7%, *p*<0.001). Some aspects of medical care differed between the groups, as only one patient with CI was admitted to the ICU, and fewer of them received non-invasive mechanical ventilation (7.1% *vs*. 25.4%, *p*<0.001). Palliative was provided more frequently in subjects with CI (79.2% *vs*. 66.3%, *p*=0.038).

**Table 2.**
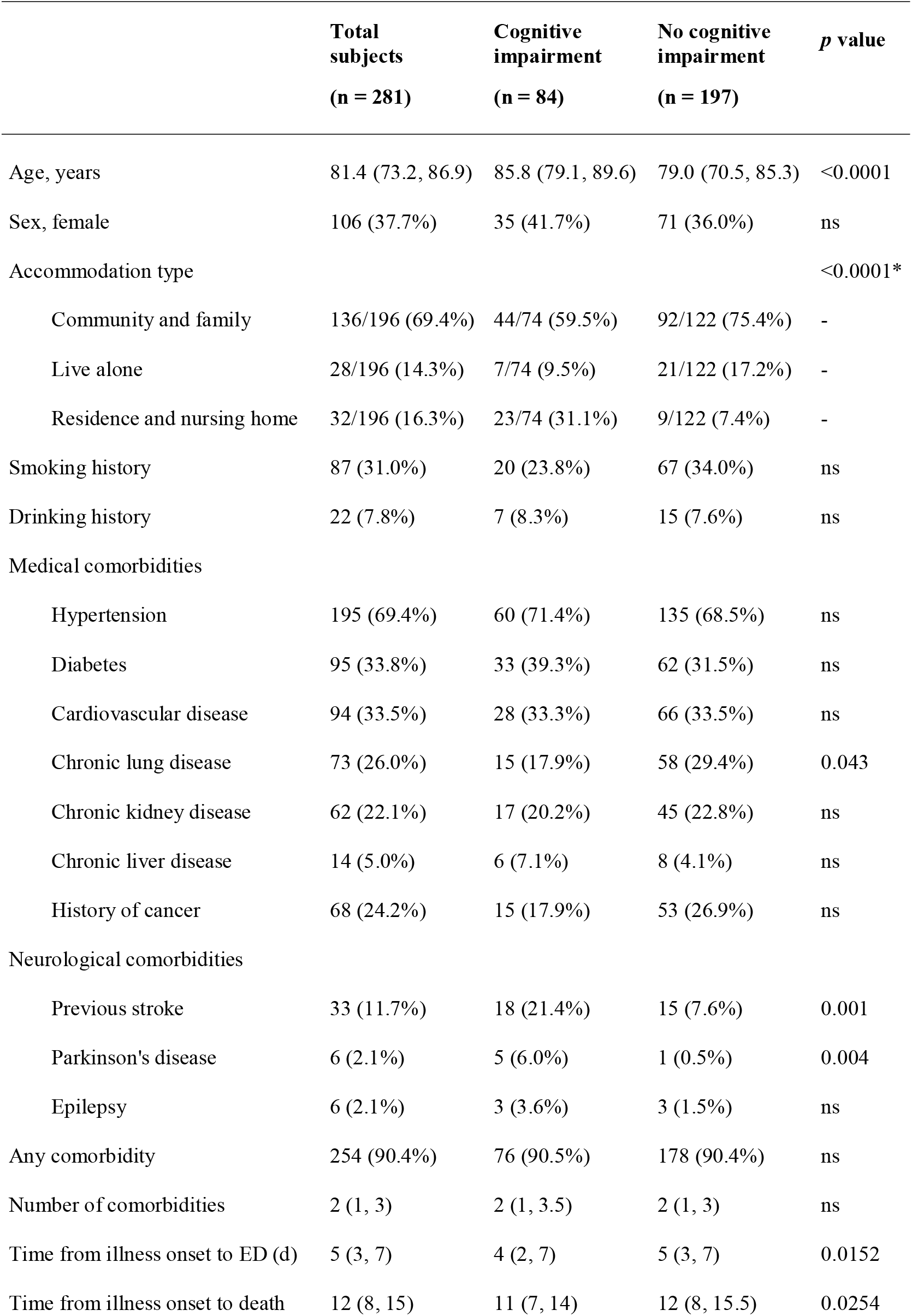
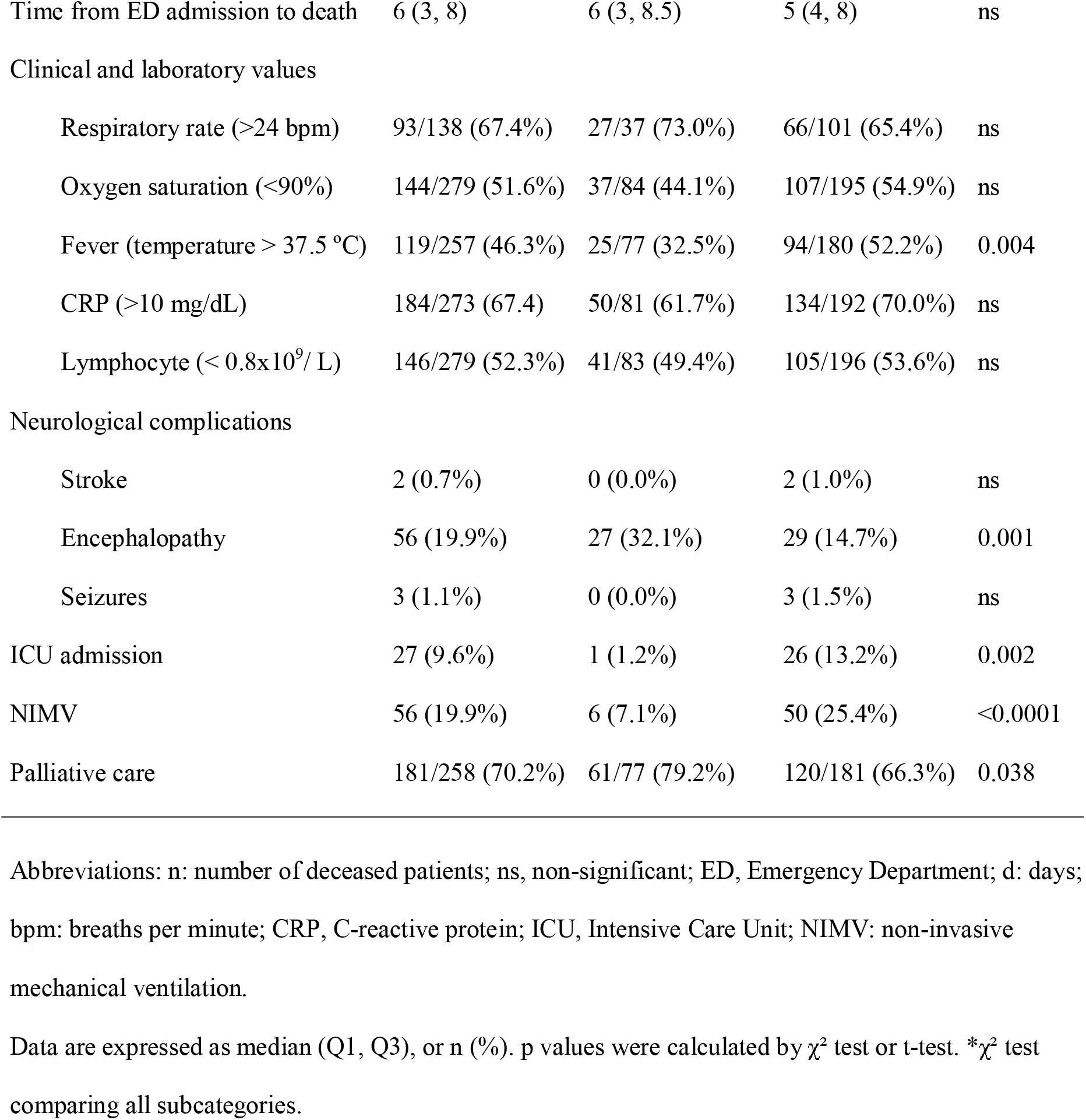
Demographic and clinical characteristics of confirmed COVID-19 deceased patients with and without cognitive impairment.

## DISCUSSION

In March 2020, 339 people have died of confirmed or suspected COVID-19 in our hospital, almost three times more deaths than all the other causes combined during the whole month. Severe COVID-19 is affecting distinctively older people with chronic diseases. We found that CI is one of the most common comorbidities in confirmed COVID-19 (30% of the subjects). This is not surprising, because dementia is a prevalent and sometimes overlooked underlying cause of death (11), and respiratory infections are one of the leading immediate cause of death in this population (12). Although the percentage of CI in COVID-19 deceased subjects is high, our results are not informative on the risk that CI has on mortality. A recent report has described that dementia could be a risk factor for mortality in COVID-19 (13), and future researhc will clarify this issue and explore possible pathophysiological mechanisms. For example, a recent report has described that the ApoE e4e4 allele increases the risk of severe COVID-19 infection, independent of pre-existing dementia (14).

As expected, medical care was somewhat different in COVID-19 patients with CI, as they were rarely admitted to the ICU and were treated more frequently with palliative care to prioritize comfort and symptom management.

Our study has some limitations. First, due to its retrospective design, information about some of the variables may be incomplete. We tried to carefully review all the clinical notes in the electronic medical records, but some diagnoses of CI may not have been recorded. Second, only fatal cases of COVID-19 were included, and we cannot analyse whether CI is a risk factor for mortality in COVID-19. Third, the study is hospital-based, and many community-based cohort studies have shown that not all the persons with dementia seek medical attention, especially those who are very old or have a lower socio-economic status (15). More importantly, health authorities and research reports are describing a high case fatality rate of COVID-19 in nursing homes (16), where people are dying without hospital transfer. By April 17, 2020, more than 20,000 people with confirmed COVID-19 had died in Spain, 7,132 of them in the Community of Madrid (17). These numbers are distressing, but they probably reflect only a part of the impact of COVID-19. The local health authorities have reported that from March 8 to April 17, 2020, 5,272 of the 44,132 nursing home residents in the Madrid region have died with symptoms of COVID-19, but the disease could only be confirmed in 837 of them (18). Considering that the estimated prevalence of dementia in nursing homes of western European countries ranges from 50 to 75% (19), the number of deaths in people with dementia during the COVID-19 pandemic will be probably high. The 30% of deceased COVID-19 patients with CI in our study could be an underestimation of the situation in the community. More research in nursing homes will be needed, taking into account that dementia is usually under-reported in death certificates (20).

To our knowledge, at least six clinical series have described the clinical characteristics of fatal cases with confirmed COVID-19 in Asian populations (4–9). These studies have not included CI in the analysis of comorbidities in COVID-19. A report described the clinical characteristics of 54 deceased COVID-19 patients (8), and included as a comorbidity the category “neurologic disease” that grouped together dementia and stroke. The proportion of patients with neurologic disease was 18.5%, with a marked difference between those under 70 years (0%), and those over 70 years (29.4%). The results in the older group could be comparable to our findings. A short report from Italy mentions a preliminary study in a subsample of 355 patients with COVID-19 who died and underwent chart review (21). They were also old (mean age 79.5 years), more frequently male, and 6.8% had dementia. It is a lower proportion than our study (21.1%), but the brief clinical description does not allow more comparisons.

In summary, our study shows that CI is a common comorbidity in COVID-19 deceased patients, and it has been relatively overlooked until now. In fact, many guidelines do not include dementia as a vulnerable group for severe COVID-19. When new public health threats like COVID-19 emerge, society, governments and research agencies should not forget previous medical priorities, like neurodegenerative diseases.

## Data Availability

Anonymized data may be available upon reasonable request from qualified researchers.

## ACKNOWLEDGMENTS

We would like to thank the members of the Scientific Support Unit (Epidemiology, Bioinformatics and Biostatistics) of the Instituto de Investigación Hospital 12 de Octubre, for their help with data extraction.

